# Impact of social vulnerability on cardiac arrest mortality in the United States, 2016-2020

**DOI:** 10.1101/2023.08.02.23293573

**Authors:** Karthik Gonuguntla, Muchi Ditah Chobufo, Ayesha Shaik, Neel Patel, Mouna Penmetsa, Yasar Sattar, Harshith Thyagaturu, Paul S. Chan, Sudarshan Balla

## Abstract

**Importance:** Cardiac arrest is one of the leading causes of morbidity and mortality, with an estimated 340,000 out-of-hospital and 292,000 in-hospital cardiac arrest events per year in the U.S. Survival rates are lower in certain racial and socioeconomic groups.

**Objective:** To examine the impact of social determinants on cardiac arrest mortality among adults stratified by age, race, and sex in the U.S.

**Design:** A county-level cross-sectional longitudinal study using death data between 2016 and 2020 from the Centers for Disease Control and Prevention’s (CDC) Wide-Ranging Online Data for Epidemiologic Research (WONDER) database.

**Setting:** Using the multiple causes of death dataset from the CDC’s WONDER database, cardiac arrests were identified using the International Classification of Diseases (ICD), tenth revision, clinical modification codes.

**Participants:** Individuals aged 15 years or more whose death was attributed to cardiac arrest.

**Exposures:** Social vulnerability index (SVI), reported by the CDC, is a composite measure that includes socioeconomic vulnerability, household composition, disability, minority status and language, and housing and transportation domains.

**Main outcomes and measures:** Cardiac arrest mortality per 100,000 adults.

**Results:** Overall age-adjusted cardiac arrest mortality (AAMR) during the study period was 95.6 per 100,000 persons. The AAMR was higher for men as compared with women (119.6 vs. 89.9 per 100,000) and for Black, as compared with White, adults (150.4 vs. 92.3 per 100,000). The AAMR increased from 64.8 per 100,000 persons in counties in Quintile 1 (Q1) of SVI to 141 per 100,000 persons in Quintile 5, with an average increase of 13% (95% CI: 9.8-16.9) in AAMR per quintile increase.

**Conclusion and relevance:** Mortality from cardiac arrest varies widely, with a more than 2-fold difference between the counties with the highest and lowest social vulnerability, highlighting the differential burden of cardiac arrest deaths throughout the U.S. based on social determinants of health.

**Key Points:** *Question:* What is the impact of social determinants of health on cardiac arrest mortality in the United States (U.S.)?

*Findings:* In this national cross-sectional study spanning 5 years, we found that counties in higher quintiles of the social vulnerability index had higher rates of mortality from cardiac arrest, which was consistent across all subgroups.

*Meaning:* Our data suggests that sizable disparities in cardiac arrest remain in the U.S. A multi- dimensional approach to address gaps, with emphasis on social determinants of health, is needed for vulnerable populations to impact outcomes.

## Introduction

Cardiac arrest is one of the leading causes of morbidity and mortality in the U.S. Out-of-hospital cardiac arrest (OHCA) accounts for over 340,000 events each year and in-hospital cardiac arrest (IHCA) is estimated to be around 292,000 events per year ^1^. Prior studies have reported sex- and racial differences in survival at the patient- and neighborhood-level from OHCA^1,2^ and IHCA ^3,4^. Other studies have identified differences in survival rates for OHCA based on neighborhood income, with lower survival rates in communities with lower median household incomes.^3^ However, most prior studies have two major shortcomings. Reports of case-survival rates for cardiac arrest ignore the fact that the incidence of cardiac arrest may also differ by race and socioeconomic status. Therefore, prior reports may have underestimated differences in cardiac arrest burden by race or socioeconomic group. Moreover, the socioeconomic factors associated with survival after cardiac arrest in prior studies likely extends beyond race and income.

In Canada and Korea, the Ontario marginalization index and the Carstairs index have been used to assess the association of multiple neighborhood characteristics on case-survival from OHCA.^5,6^ In the U.S., the Social Vulnerability Index (SVI) provides multiple dimensions on social vulnerability beyond race and income. Developed by the Centers for Disease Control and Prevention (CDC), the SVI is a comprehensive county-level measure encompassing most social determinants of health indicators. ^5–7^ The SVI incorporates 15 social factors from four main domains (socioeconomic status, household composition, race/ethnicity/language, and housing/ transportation) to create a vulnerability score between 0 (least vulnerable) to 1(most vulnerable) to identify communities with a differential burden of disease beyond using individual racial and socioeconomic variables (Figure 1).

**Figure 1:**
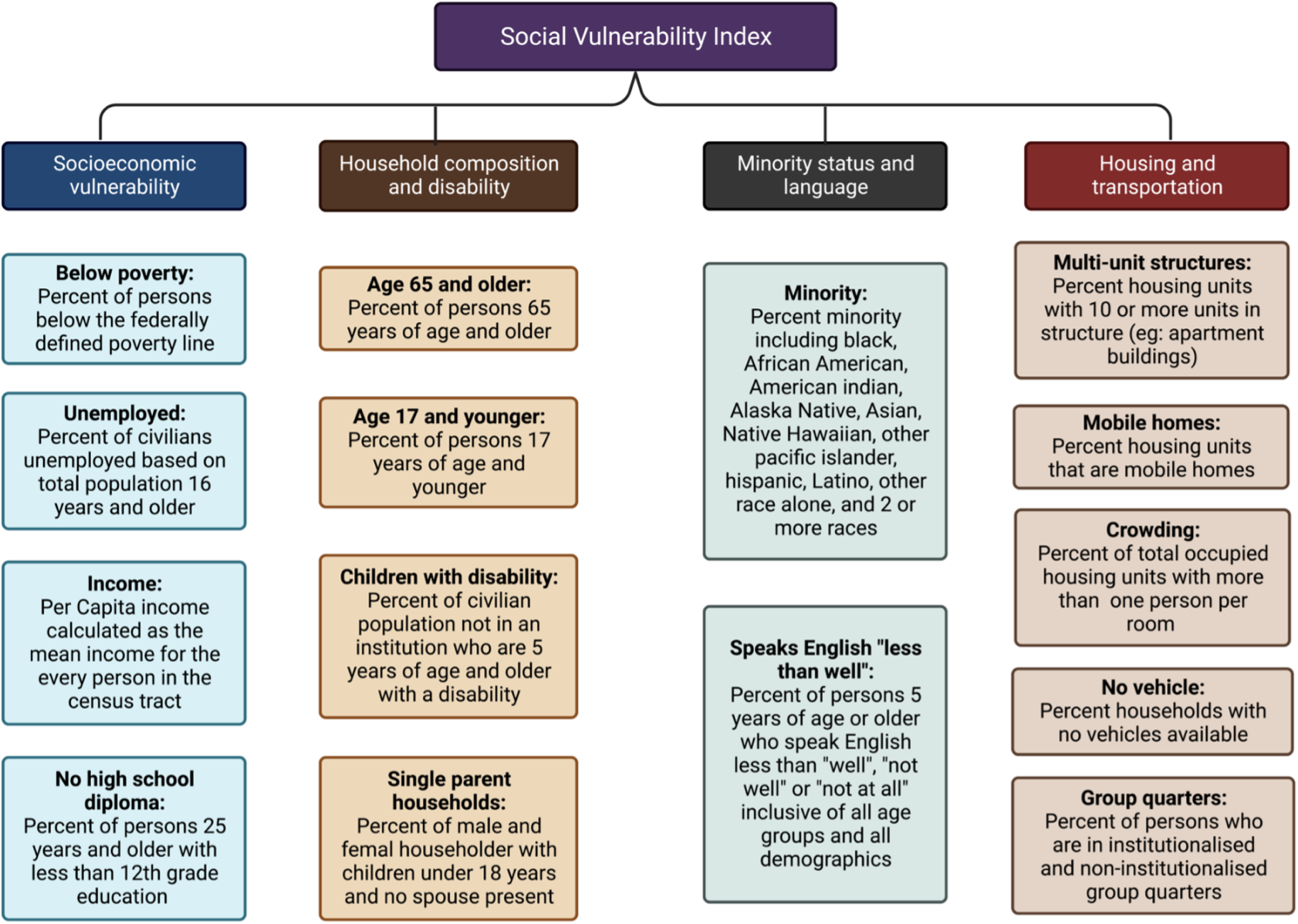
Visual representation of various components of the social vulnerability index.

Accordingly, we leveraged U.S. mortality data linked with county-level SVI data to examine cardiac arrest mortality rates. We then assessed the association between county-level SVI (a community measure of its social determinants of health) and cardiac arrest mortality.

## Methods

We utilized the Centers for Disease Control and Prevention’s Wide-Ranging Online Data for Epidemiologic Research (CDC WONDER) multiple causes of death dataset to analyse deaths occurring in the U.S.^8–10^ This data is based on death certificates, and each death certificate contains a single underlying cause of death and up to 20 additional multiple causes, as well as patient level sociodemographic data. The underlying cause of death is defined as the “disease or injury which initiated the train of events leading directly to death.”

For this study, we included data on all adults 15 years of age or older between January 1, 2016, and December 31, 2020, in the CDC-WONDER database with cardiac arrest listed as a contributing or underlying cause of death using International Statistical Classification of Diseases, Tenth Revision, (ICD-10) code I46 (I46.0, I46.1, I46.9). Age-adjusted mortality rate and corresponding standard errors for all deaths with cardiac arrests listed as the underlying or contributing cause of death were obtained using the death data from 2016 to 2020 for the entire U.S. We calculated the AAMRs by standardizing the cardiac arrest related deaths to the 2000 U.S population^9^. Race and ethnicity were assessed in accordance with standards from the U.S. Office of Management and Budget. Ethnicity was categorized as Hispanic, non-Hispanic Black/African American, or non-Hispanic White. Race was classified as American Indian or Alaskan Native, Asian/Pacific Islander, Black or African American, and White.

### Social Vulnerability Index (SVI)

Every U.S. County and census tract’s relative vulnerability are outlined by CDC’s Agency for Toxic Substances and Disease Registry. ^11^ The SVI ranges from 0 to 1 and is comprised of 4 components with 15 U.S census variables to help officials identify groups who are more susceptible during pandemics and disasters: 1) socioeconomic status (below poverty, unemployed, income level, and no high school diploma); 2) household composition and disability (aged 65 years or older, aged 17 years or younger, individuals >5 years with a disability, and single-parent households); 3) minority status and language (minority and individual speaks English “less than well”); and 4) housing and transportation (multiunit structure, mobile home, crowding, group quarters, and no vehicle). Each of the 15 census-level variables are ranked from 0 to 1, with higher values exhibiting greater vulnerability. SVI was classified into five quintiles (Q1-Q5).

## Statistical analysis

Counties were classified into five quintiles based on their SVI, with SVI-Q1 being the least vulnerable and SVI-Q5 being the most vulnerable. We aggregated counties across SVI quintiles to compare AAMRs among quintiles. Counties with <20 deaths were excluded from analysis because data were excluded due to low volumes and likely sparse populations. Using county-reported AAMR, we computed the mean AAMR, standard errors (SE), and 95% confidence intervals (CI). The Joinpoint regression program (version 4.9.1.0; National Cancer Institute) was used to describe trends in mortality rates. ^12^ We created maps utilizing AAMR per 100,000 by SVI quintiles. County-level AAMRs per 100,000 person-years with 95% CIs were estimated for the overall population and stratified by demographic variables (age, sex, race, and ethnicity) and urbanization. Continuous variables (e.g., county attributes used in the SVI) are reported as medians (IQR).

According to the 2013 U.S. census classification, National Center for Health Statistics Urban-Rural Classification Scheme (NCHS) was used to divide the entire population based on its size into urban (large metropolitan area [population, ≥1 million], medium/small metropolitan area [population, 50,000-999,999]), and rural (population, <50,000) counties. ^6,13^ Metropolitan areas were grouped as large metro (large central metro, large fringe metro), small metro (medium metro, small metro), and non-metro areas (micropolitan and non-core). We further stratified the age-adjusted cardiac arrest mortality rate (AAMR) by age, sex, race/ethnicity, census region, state, and urban-rural classification and assessed the impact of SVI on cardiac arrest mortality. The study was exempt from our institutional review board approval, given the deidentified nature of the database.

## Results

Of the 3,148 counties, 2,876 met criteria for calculations of AAMR and comprised the study cohort. Overall, there were 1,521,938 cardiac arrest-related deaths in U.S. counties between 2016 and 2020. The mean AAMR was 95.6 per 100,000 persons (95% CI: 95.4 – 95.7) and ranged from 6.01 (Olmsted county, MN) to 768.12 (Alcorn county, MS). Following a steady decline from 2016 (94.5 per 100,000 persons [95% CI: 94.2- 94.8]) through 2019 (89.7 per 100,000 persons [95% CI: 89.4–90.0]), there was a sharp rise in 2020 to 104.5 per 100,000 persons (95% CI: 104.2-104.8). The AAMR for Black adults was 150.4 per 100,000 persons as compared with 92.3 per 100,000 persons for White adults (Table 1). The AAMR was 89.9 per 100,000 persons for women as compared with 119.6 per 100,000 persons for men. The AAMR was highest in non-metropolitan areas (104.2 per 100,000 persons) and lowest in medium and small metropolitan areas (85.6 per 100,000 persons).

**Table 1:** AAMR (per 100,000 residents) by select sociodemographic characteristics in the different quintiles of SVI.

| Variable | Category | Overall | Quintile<br>1 | Quintile<br>2 | Quintile<br>3 | Quintile<br>4 | Quintile<br>5 |
| --- | --- | --- | --- | --- | --- | --- | --- |
| All |  | 95.6 | 64.8 | 78.3 | 90.9 | 105.4 | 141.1 |
| Sex | Male | 119.6 | 79.7 | 95.4 | 111.4 | 126.4 | 171.9 |
|  | Female | 89.9 | 54.2 | 66.4 | 78.1 | 88.5 | 124.8 |
| Age | <65 | 35.8 | 17.3 | 23.8 | 30.2 | 39.5 | 55.3 |
|  | >=65 | 549.7 | 392.7 | 455.0 | 525.0 | 570.1 | 766.6 |
| Race/Ethnicity |  |  |  |  |  |  |  |
|  | White | 92.3 | 64.3 | 75.8 | 88.1 | 99.7 | 129.2 |
|  | Blacks | 150.4 | 95.7 | 123.2 | 149.2 | 173.0 | 209.0 |
|  | Hispanics | 97.5 | 64.9 | 76.9 | 90.4 | 104.7 | 145.2 |
|  | Asian/Pacific<br>Islander | 68.5 | 47.1 | 53.5 | 68.8 | 67.5 | 97.5 |
|  | American<br>native | 175.0 | 94.0 | 114.4 | 144.4 | 153.3 | 205.9 |
|  | Others | 150.5 | 70.3 | 98.6 | 127.3 | 154.9 | 196.4 |
| Urbanization |  |  |  |  |  |  |  |
|  | Large | 95.8 | 67.5 | 93.7 | 99.9 | 92.8 | 125.2 |
|  | Medium/Small | 85.6 | 58.6 | 67.6 | 82.7 | 97.9 | 119.8 |
|  | Non-Metro | 104.2 | 69.7 | 81.8 | 90.6 | 115.1 | 144.2 |

The distribution of SVI by quintiles is shown in Figure 2. For counties in the lowest social vulnerability (Q1 of SVI), the AAMR was 64.8±1.8 per 100,000 persons. In contrast, counties with the highest social vulnerability (Q5 of SVI) had an AAMR of 141±3.9 per 100,000 persons, with an average increase of 13% (95% CI: 9.8-16.9) deaths per quintile increase (Figure 3). This was consistent across the 4 components of the total SVI on sensitivity analysis.

**Figure 2:**
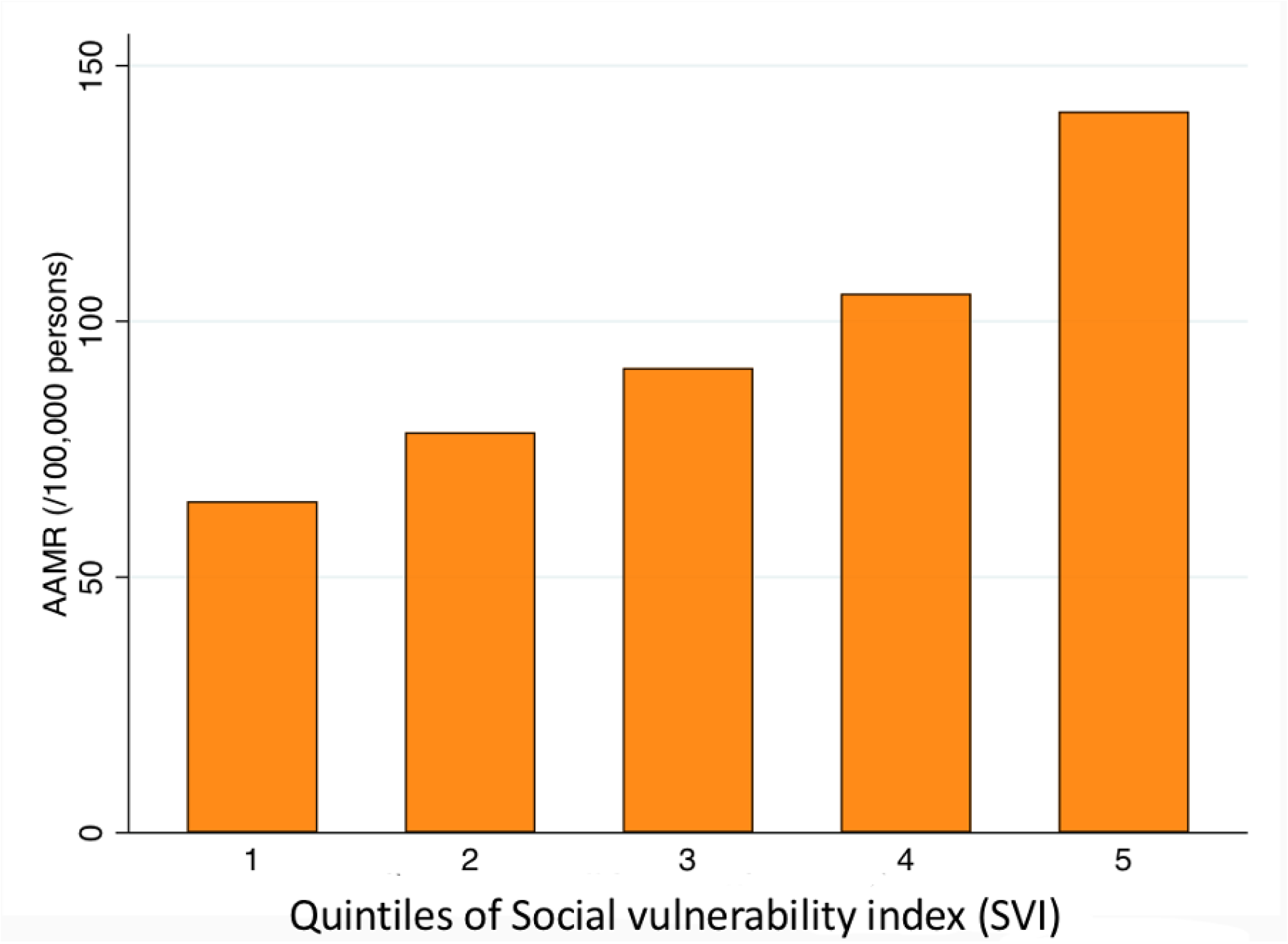
AAMR by quintiles of social vulnerability index in the USA: 2016-2020.

**Figure 3:**
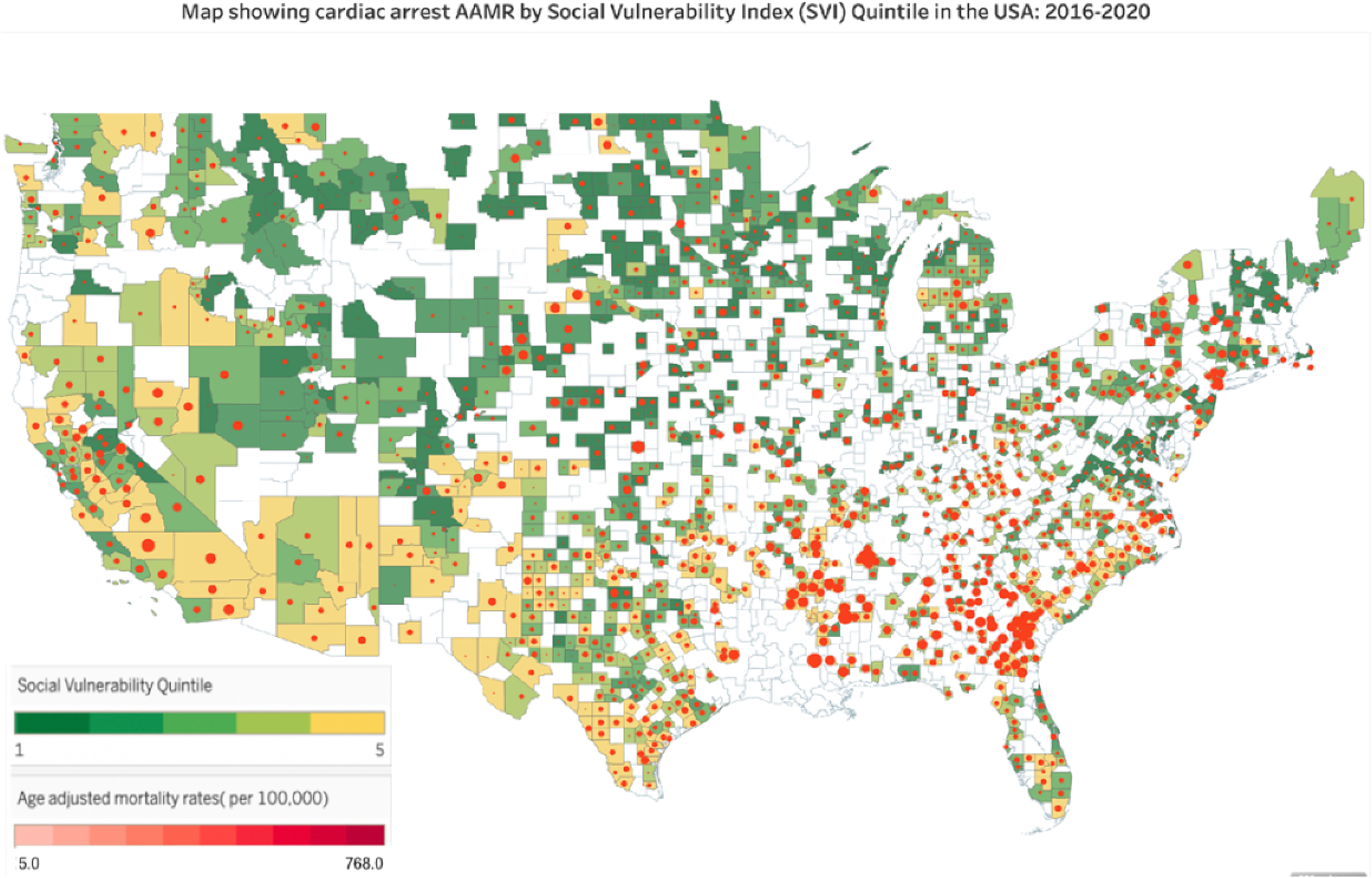
Map showing cardiac arrests related mortality by social vulnerability quintiles in the USA: 2016-2020

## Discussion

In our study using death certificate data from the CDC WONDER database, there was an association between county-level SVI and AAMR from cardiac arrest, with increasing mortality in the most vulnerable counties in higher quintiles of SVI. The increase in mortality rate in counties with higher SVI was consistent across age, sex, race and ethnicity, and urbanization subgroups.

The association of race and socioeconomic status with mortality from cardiac arrest has been described previously^3,14,15^. Data from the Cardiac Arrest Registry to Enhance Survival registry have found lower survival rates in majority Black and non-high income neighborhoods^3,16^. A similar association between cardiac arrest outcomes and a few social determinants has been demonstrated for IHCA as well. Black race, low individual income, and low household assets have been associated with worse outcomes in IHCA^4,17–20^. A Swedish study found that patients with lower socioeconomic status based on income and education had worse outcomes from IHCA^21^. Our study extends the results of previously published papers but differs in a few aspects. First, our study incorporates all cardiac arrest deaths, including IHCA and OHCA. Second, we examine the burden of death due to cardiac arrest in the community rather than the survival rates among those with cardiac arrest. Third and novel to our study is the impact of social determinants of health on cardiac arrest mortality in the U.S. using SVI as a measure of social determinants of health at the county level. SVI includes domains other than those that have been studied in prior studies. For example, housing characteristics are included in the SVI and are also likely to impact outcomes in CA. Indeed, Drennan et al. reported lower survival rates in patients suffering OHCA residing on higher floors in high-rise apartment buildings. ^22^ Last, the association of county-level SVI with CA mortality and the readily available nature of SVI data could provide a framework to identify vulnerable counties to institute policy changes and resource allocation.

Geographic disparities in cardiac arrest mortality at the county level were observed, with counties in higher SVI quintiles (most vulnerable) having worse mortality rates compared to lower SVI quintiles (least vulnerable). Further, for each quintile increase in SVI, there was a 13% increase in AAMR. It is also noteworthy that counties in higher quintiles of SVI had higher AAMR, irrespective of their age, sex, and race. The higher mortality in vulnerable counties is likely from a multitude of factors. Social determinants play a crucial role in factors linked to the chain of survival after cardiac arrest. Furthermore, underlying health conditions and recognition of symptoms prior to cardiac arrest are also impacted by social determinants.

Bystander CPR has been shown to be crucial in improving survival after cardiac arrest. Several variables included in the SVI have shown an association with rates of bystander CPR.^23–25^ In fact, neighborhood characteristics such as median household income, percentage of people living below the poverty line, percentage of single-person households, racial or ethnic-group composition, and percentage of people with a high-school diploma or a higher level of education have been associated with lower rates of bystander CPR^26–28^. The study by Sasson et al showed a 50% lower likelihood of bystander CPR in low-income Black neighborhoods compared to high-income non-Black neighborhoods.^2^ Second, patients with lower socioeconomic status have been found to have a higher prevalence of non-monitored/unwitnessed arrests, with mortality being higher in this cohort.^29^ Third, the incidence of cardiac arrest has been associated with social determinants. Black race, low education attainment, low socio-economic status, with lower household and personal income are associated with a higher incidence of OHCA as well as sudden cardiac deaths, Fourth, poor health conditions are also associated with lower education level, and unemployed status.^30–34^ Risk factors for heart disease, such as smoking, cholesterol, hypertension and diabetes, may be suboptimal and predispose to sudden death. Indeed, an inverse gradient has been reported in cohorts from the United States and Finland between the above risk factors and socioeconomic status.^32^ Last, social determinants of health have been associated with the under-recognition of symptoms for potentially fatal cardiovascular conditions, such as myocardial infarction and stroke.^35–37^

System-level factors that impact CA mortality, such as disparities in CPR training, have also been reported thus far. Significant variability has been reported across counties with regard to CPR training which is likely to affect rates of bystander CPR. Variables associated with lower rates of CPR training in counties mirror those associated with low bystander CPR rates, including increasing proportions of residents living in rural areas, high proportions of Black or Hispanic residents, lower median household income levels, and fewer physicians. Timely activation and the arrival of EMS could also be an issue in vulnerable counties^14^. SES has also been associated with shorter out-of-hospital transport intervals ^38^ Furthermore, in hospital settings, the Black race was found to be associated with a delay in defibrillation after IHCA. ^3,39^

Potential barriers to improving outcomes in these counties include access to CPR training and mitigating linguistic barriers due to a lack of English language proficiency. Education about the prevention of atherosclerotic cardiovascular diseases and symptoms of myocardial infarction, stroke, and cardiac arrest could also be initiated in vulnerable counties. A set of targeted interventions in the vulnerable counties that address the entire chain of survival in OHCA is the need of the hour to resolve inequities. A novel concept of using a drone to deliver AED for OHCA has been reported (38). Considering that smartphones are currently ubiquitous, a recent study evaluated the use of smartphone dispatch of volunteers to the site of out-of-hospital cardiac arrest. ^40^ Whether the implementation of such novel strategies will mitigate socioeconomic disparities in cardiac arrest mortality requires further study.

## Limitations

Even though the CDC WONDER is a large database, there are some limitations to the study due to shortcomings of the database. First, the dataset is obtained from death certificates and relies on the accuracy of data in the death certificate. Moreover, the data does not include information about co-morbidities, prior medical and surgical history, medications, vital signs, laboratory information, treatment, and information about CPR and AED utilization. Second, cardiac arrest in our cohort included both out-of-hospital and in-hospital cardiac arrest and cannot be differentiated in the dataset. Similarly, the etiology of cardiac arrest– traumatic vs. non-traumatic cannot be discerned. Third, we performed a retrospective cross-sectional analysis, so we were not able to prove causal relationships between county-level characteristics and mortality. Fourth, individual risk factors could not be examined, so we performed crude estimates of AAMR. The database provides us with aggregated data and not individual-level data, which limits our ability to perform adjusted analyses. To control these limitations, we performed a subgroup analysis looking at demographic and population-level variables. Finally, SVI was used to assess social vulnerability, which is a comprehensive tool to assess social factors which can predispose a population to various disadvantages. However, SVI does not include various other important variables such as food insecurity, community and social contextual factors, and healthcare access barriers such as proximity to quality healthcare center and insurance status, among others.

## Conclusion

Social determinants of health as assessed by the SVI were associated with cardiac arrest mortality, with vulnerable counties in higher SVI quintiles having higher overall cardiac arrest mortality. The increased mortality rate in vulnerable counties was noted across different age, sex, and racial subgroups and metropolitan status. A multi-dimensional approach with an emphasis on social determinants of health could allow for targeted interventions to address the higher cardiac arrest mortality burden in vulnerable counties.

## Visual abstract

Visual representation of the impact of the social vulnerability index on cardiac arrest mortality ^41^

## Data Availability

Data derived from public domain resources

https://wonder.cdc.gov/

---

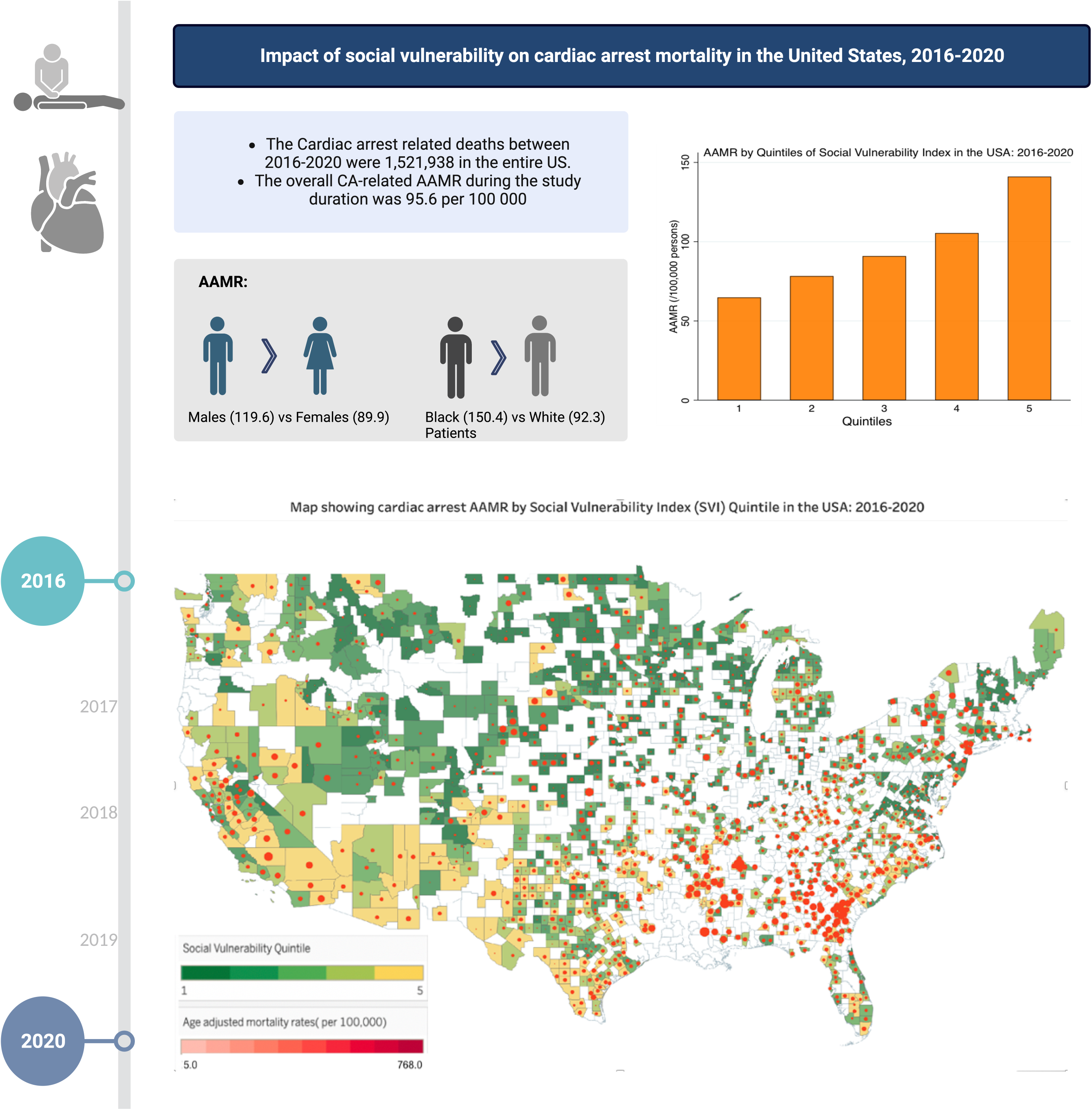

